# The Norwegian Microbiota Study in Anorexia Nervosa (NORMA): integrating a clinical trial with preclinical experiments – a study protocol

**DOI:** 10.64898/2026.01.21.26344578

**Authors:** Ida Heir Hovland, Lasse Bang, Anne Mari Herfindal, Stine Sofie Strømland, Tina Bogetvedt Spernes, Armita Jahanshahi, Kathinka Larsen Otterdal, Dunja Arsenovic, Trude Elise Aspholm, Ylva Vik, Jenny HM Storvik, Monica Hauger Carlsen, Monica Linnea Ones, Renata Alisauskiene, Kristina Hansen, Siri Weider, Ivan Samdal, Johan Dahl, Hilde Tveit Reistad, Åsne Skram Trømborg, Lars Jordhøy Lindstad, Signe Birkeland, Hanne Torp Eriksen, Jeanette Engeset, Cynthia M. Bulik, Bjørge Westereng, Harald Carlsen, Øyvind Rø, Siv Kjølsrud Bøhn

**Affiliations:** Faculty of Chemistry, Biotechnology and Food Sciences, Norwegian University of Life Sciences, Ås, Norway; Department of Child Health and Development, Norwegian Institute of Public Health, Oslo, Norway; The Regional Department for Eating Disorders, Oslo University Hospital, Oslo, Norway; Department of Nutrition, Institute of Basic Medical Sciences, University of Oslo, Oslo, Norway; The regional Department of Eating Disorders, Haukeland University Hospital, Bergen, Norway; Regional Center for Eating Disorder, Nordland Hospital, Bodø, Norway; Regional Eating Disorder Unit, Nord-Trøndelag Hospital Trust, Levanger, Norway; Research institute of Modum Bad, Vikersund, Norway; District Psychiatric Center, Innlandet Hospital, Gjøvik, Norway; District Psychiatric Center, Søndre Oslo, Norway; Department of Medical Epidemiology and Biostatistics, Karolinska Institutet, Stockholm, Sweden; Department of Psychiatry, University of North Carolina at Chapel Hill, Chapel Hill, USA; Department of Nutrition, University of North Carolina at Chapel Hill, Chapel Hill, USA; Institute of Clinical Medicine, University of Oslo, Oslo, Norway; Department of Psychology, the Norwegian University of Science and Technology (NTNU), Trondheim, Norway

**Keywords:** anorexia nervosa, anxiety, gut-brain axis, clinical study, gastrointestinal symptoms, diet, prebiotics, in vitro fermentation, mouse study, fecal microbiota transplantation

## Abstract

**Background:** Anorexia nervosa (AN) remains difficult to treat, partly due to co-occurring mental health challenges and gastrointestinal symptoms. Emerging research suggests that individuals with AN exhibit gut microbiota dysbiosis and dysregulation in the gut-brain axis (GBA). However, research examining the role of gut microbiota as a potential driver of AN-related pathologies remains limited. The Norwegian Microbiota Study in Anorexia Nervosa (NORMA) will therefore investigate gut microbiota and its interaction with the GBA in AN.

**Methods:** NORMA is a collaboration between the Norwegian University of Life Sciences and seven Norwegian specialized eating disorder inpatient treatment units, consisting of three work packages (WP): a clinical observational trial (WP1), in vitro fermentation experiments (WP2), and animal experiments (WP3). In WP1, 90 patients with AN (age 16-50, BMI<18.5) admitted for treatment and 90 healthy controls (HCs, age 16-50, BMI 18.5-27) will be recruited. Data on mental and physical health, dietary intake, and blood and fecal samples for biomarker and microbiota analyses will be collected at baseline, 6 and 12 weeks after start of treatment for AN patients and once for HCs. Outcomes will be compared between groups, and longitudinal effects of standard treatment examined within the AN group. In WP2, fecal microorganisms from patients and HCs will be grown *in vitro* to assess influence of prebiotics. In WP3, mice will receive fecal microbiota from AN and HC donors to determine if and how AN-related microbiota affects AN-relevant phenotypes.

**Conclusion:** NORMA is pioneering in its integration of clinical, *in vitro,* and animal studies, providing the most comprehensive gut microbiota study of AN so far. By investigating the role of gut microbiota in AN and effects of standardized treatment on gut microbiota composition, this study aims to inform the development of innovative therapeutic strategies and ultimately improve treatment outcomes and life quality for individuals with AN.

**Trial registration:** NORMA is a registered clinical trial: clinicaltrials.gov as NCT06144905.

## Background

Anorexia nervosa (AN) is a severe eating disorder (ED) associated with high rates of medical complications, psychiatric comorbidities, and mortality (1, 2). The lifetime prevalence of AN in Europe is estimated to be 1-4% (3). Globally, the AN prevalence is rising, especially among adolescents, and the COVID-19 pandemic may have further increased both the prevalence and severity of the condition (4). In Norway, approximately 3000 females may require treatment for AN at any given time (5), and the incidence of AN in specialized health care services has increased post pandemic (6). The etiology of AN remains elusive, yet it is clear that a combination of genetic (7) and environmental factors (8) significantly contributes to its development, with several risk factors identified (9). Patients with AN often struggle with intense fear of weight gain, low weight, and ambivalence toward recovery, challenges that are compounded by significant gastrointestinal (GI) symptoms (10, 11), reported in up to 90% of patients (12). These factors may complicate treatment, leading to high dropout-rates (13), and present challenges not only to the patients, but also their families, clinicians and society at large, due to the high societal costs involved (14).

Currently, no gold-standard treatment exists for patients with AN. However, specialized care typically combines psychotherapy, nutritional therapy, and management of co-occurring complications and comorbidities. Unfortunately, treatment outcomes are often unsatisfactory (15), with approximately 50% of patients experiencing poor recovery and 20% developing a protracted course (16). Despite its severity, current dietary interventions are largely based on clinical experience and general guidelines, rather than evidence-based treatments (17), underscoring the need for more effective treatment alternatives. In recent decades, there has been increased focus on the role of gut microbiota in AN, both in its potential involvement in pathogenesis (18) and as a promising therapeutic target (19).

The human microbiota refers to the vast collection of microorganisms that colonize the human body, whose genomes encode more than three million genes that play a significant role in both physical and mental health (20). The majority of these microbes reside in the GI tract and include bacteria, viruses, fungi, and other microbes (20). Although the gut microbiota has a heritable component and is affected by age, its composition is mainly influenced by environmental factors, with diet being the most potent modulator (20, 21). Patients with AN often display abnormal dietary patterns, including low energy intake, limited food variety, and fat avoidance (22, 23), and patients undergo substantial nutritional changes during treatment. Indeed, substantial evidence reveals that the gut microbiota in individuals with AN is different from healthy controls (HCs), affecting both microbial composition and function (18, 24–27). These differences are often referred to as gut microbiota dysbiosis or imbalance (2, 19).

The gut microbiota in individuals with AN has been found to differ from that of HCs in several ways. Multiple studies have reported reduced bacterial richness and evenness in AN, referred to as alpha diversity (19, 27–29), with one study showing correlation between reduced bacterial richness and depression as well as core ED psychopathology (28). However, other studies have found no significant differences in diversity indices between AN and HCs (24–26, 30), and findings related to beta diversity are similarly inconclusive (19). Thus, while there is broad evidence of and compositional alterations (i.e., gut microbiota dysbiosis) in patients with AN compared to HCs (18, 24–27), Scala et al. (19) highlight the lack of consensus concerning diversity and psychopathological symptoms in AN.

Due to the contradictory findings on bacterial alpha– and beta diversity measures, recent research on gut microbiota in individuals with AN has shifted the focus toward the investigation of specific microbial taxa (19). For instance, reduced abundances of the bacterial genera *Roseburia*, *Lactobacillus*, *Faecalibacterium*, *Clostridium*, and *Bifidobacterium* have been associated with anxiety, depression, and AN-related psychopathology (19). High abundance of the archaeon *Methanobrevibacter smithii* has also been observed in AN (24, 30, 31), a specie thought to contribute to constipation, a common GI symptom in this population (11). Additionally, higher abundances of the genus *Sutterella* have been linked to greater weight gain at one year-follow up after treatment, suggesting its potential as a prognostic marker and therapeutic target (26). Several studies have further associated gut microbiota dysbiosis with AN disease severity (18, 28, 29), possibly reflecting a metabolically dysfunctional microbial profile (32). Nevertheless, the specific features of microbial alterations in AN remain unclear due to limited sample sizes, methodological differences, and contradictory findings across studies (19, 33).

Particularly, the extent to which gut microbiota dysbiosis in AN normalizes or persists following weight restoration remains understudied, primarily due to the limited number of longitudinal studies that follow the patients throughout their treatment programs (19, 33). Moreover, the existing longitudinal studies often involve short follow-up periods, and many patients remain underweight at the conclusion of these studies, making it difficult to distinguish microbiota changes associated with underweight from those specific to AN. To date, Andreani et al. (26) have conducted the only study with a one-year follow-up, revealing persistent alterations in the gut microbiota composition throughout treatment. Furthermore, even one year after treatment and following weight restoration, the microbiota profile remained partially distinct from that of HCs. These findings align with other longitudinal studies with shorter follow-up durations, which also indicate a partial shift toward normalization during treatment, yet continued divergence from HCs persists (11, 24–26, 32, 34).

The persistent differences in gut microbiota between individuals with AN compared to HCs might imply that the gut microbiota alterations play a causal role in the pathogenesis of AN and/or reinforcing the pathogenic events, rather than being merely a consequence of undernutrition and low body weight. To investigate this hypothesis mechanistically, researchers have turned to animal experiments where gut microbiota is transferred from humans to model organisms (18, 35–39), in a procedure known as fecal microbiota transplantation (FMT). Notably, recent studies have demonstrated that FMT from patients with AN and HCs to mice can, to some extent, reproduce AN-relevant behaviors such as anxiety– and obsessive-compulsive-like behaviors (36, 38). Additionally, it has been shown that AN-FMT recipient mice (18), or their offsprings (36), exhibit impaired body weight development compared to HC-FMT recipient mice. In addition to anxiety-like behavior, a recent study found that germ-free mice receiving FMT from AN patients also showed reduced food intake, increased physical activity, and elevated inflammatory responses (39). However, these findings are not universally replicated. For example, Glenny et al. (35) reported no significant differences in body weight changes or body composition in mice receiving FMT from AN donors, attributing the lack of impact to methodological variations and the limited number of human FMT donors (35). Others have suggested that incomplete microbial transfer from humans to mice also may contribute to these findings (40). Furthermore, Kooij et al. (37) did not find effects on anxiety-like behavior in AN-FMT experiments with rats. Interestingly, FMT from AN donors has been found to change the expression of endocrine biomarkers, including gene expression of hypothalamic appetite suppressors and hunger hormone (18), and protein expression of the key satiety and appetite regulators peptide YY and leptin (38). In sum, the evidence for a causal role of gut microbes in AN remains limited, primarily due to the small number of mechanistic animal studies (18, 35–38).

The specific mechanisms by which gut microbes could influence metabolic and emotional regulation in AN remain incompletely understood. However, the gut-brain axis (GBA), the bidirectional communication network involving neural, hormonal, metabolic, and immune pathways, is believed to play a central role (41). Microbial metabolites, particularly short-chain fatty acids (SCFAs) and neurotransmitters, have gained attention as potential key modulators of the GBA. SCFAs, especially butyrate, help maintain gut barrier integrity and modulate immune responses (42). Individuals with AN show lower fecal SCFA concentrations than HCs, along with reduced abundances of SCFA-producing bacteria like *Roseburia* (11, 24, 30, 43), which have been associated with increased anxiety and depression (30). The reduced SCFA levels may contribute to a low-grade proinflammatory state in the gut, increasing intestinal permeability (44). In turn, increased permeability can allow microbial metabolites to enter underlying tissues and the bloodstream, potentially triggering proinflammatory cytokines linked to mood disturbances (19) and possibly contributing to GI symptoms (45). Although there is limited support from human studies that gut permeability is increased in AN (46), one animal study has suggested a role for gut barrier dysfunction in activity-based AN (47). Furthermore, low fecal concentrations of the neurotransmitters serotonin, dopamine, and gamma-aminobutyric acid (GABA) have been reported in individuals with AN compared to HCs, supporting a possible link between microbial imbalance and neurotransmitter deficits (24). Similarly, serum bacterial metabolites in AN may mediate microbiota-related disruptions in appetite, emotion, and behavior regulation (18). While support from more mechanistic studies is needed, these findings may indicate that gut microbiota dysbiosis seen in AN could contribute to impaired neurotransmitter production, potentially contributing to emotional dysregulation.

Despite growing evidence from both animal and human studies suggesting that the gut microbiota may play a role in AN development and persistence, findings are inconsistent, and the underlying mechanisms remain unclear. This knowledge gap highlights the need for further investigation into the potential causal role of gut microbiota dysbiosis in the onset and persistence of AN, which seems increasingly likely (26). Furthermore, longitudinal studies in patients with AN during inpatient treatment are scarce and rarely include comprehensive dietary data (19, 33), despite diet being central to both AN pathology and gut microbiota composition. Consequently, the impact of AN-specific dietary patterns (e.g., energy restriction, low food diversity, fat avoidance) on the gut microbiota is largely unexplored (22, 23, 48). Additionally, gut microbiota changes during and after treatment and weight restoration are poorly understood, partly due to short follow-up periods and patients remaining underweight. Moreover, while some studies have examined the effects of probiotic supplementation on gut microbiota and AN-related pathologies (49, 50) prebiotic interventions in AN have yet to be examined (51).

Addressing these gaps is essential for developing personalized, evidence-based nutritional strategies that integrate microbiome modulation into the broader therapeutic framework for AN. To advance the field, larger longitudinal human studies with detailed dietary data and standardized animal models using a larger number of FMT donors are needed. Expanding microbiota research in AN may open new avenues for adjunctive therapies to support psychological recovery, ease GI symptoms, reduce inflammation, and restore nutritional status.

## The Norwegian Microbiota Study in Anorexia Nervosa (NORMA)

The Norwegian Microbiota Study in Anorexia Nervosa (‘the NORMA study’), consisting of three work packages (WP), is the first to combine a longitudinal, clinical observational trial (WP1), in vitro experiments (WP2), and animal experiments in mice (WP3) to investigate the role of gut microbiota in AN. This comprehensive approach is uniquely positioned to address key gaps in literature and advance our understanding of the etiology of AN, as well as the development of microbiota-informed treatments.

The primary aims of the clinical observational trial (WP1) are to identify differences in the gut microbial composition between patients with AN and HCs and to investigate whether standard clinical inpatient treatment with re-nutrition over 12 weeks will have effects on the gut microbiota. Additionally, we will examine the role of gut microbiota for GI problems, gut inflammation and permeability, and mental health outcomes such as anxiety, obsessive symptoms, and depression. Furthermore, we aim to explore how dietary factors interact with gut microbiota to affect these outcomes.

While the clinical trial can provide knowledge on the associations between the gut microbiota and AN features and clinical presentation, the in vitro (WP2) and animal (WP3) experiments will study causation between the AN microbiota and AN-relevant phenotypes and seek to identify prebiotic treatments to normalize gut microbiota dysbiosis in AN. One candidate prebiotic that will be tested is the fiber galactoglucomannan which has been shown to increase the abundance of the SCFA-producing bacteria *Roseburia* in pigs (52), though other candidates will also be evaluated. Because patients with AN often have an extreme fear of calories and rigid dietary patterns, it would be challenging to conduct dietary interventions to test new prebiotic candidates. Thus, the in vitro experimental procedure enables a much easier platform for testing multiple promising prebiotic candidates for targeted treatment of AN to optimize the gut microbiota for tolerable weight-restoration and improved health. The animal experiments will allow for in vivo investigations of human AN-microbiota transplantation in mice on gut-relevant and other AN-relevant features (e.g., appetite, weight development, anxiety– and compulsive-like behaviors) and the impact of candidate prebiotics to treat the AN phenotype.

The long-term goal of the NORMA study is to provide knowledge on the associations between the gut microbiota and AN features and clinical presentation and lay foundations for more specific dietary approaches tailored to AN. Furthermore, we aim to provide the groundwork needed to develop a personalized pharmaceutical solution to treat microbiota dysbiosis in AN. The creation of a non-energy-yielding pharmaceutical supplement to restore the gut microbiota would not only be a feasible clinical approach for AN treatment, but also hold innovative potential in the global pharmaceutical market.

By integrating clinical, experimental, and mechanistic approaches, the NORMA study aims to advance our understanding of the gut microbiota and its interaction with GBA in AN. Ultimately, this research could pave the way for personalized, microbiota-based treatment strategies that improve both physical and psychological recovery in individuals with AN.

## Methods

### The NORMA study

The NORMA study consists of WP1; a clinical observational trial (**Fig 1**), WP2; in vitro experiments (**Fig 2**), and WP3; animal experiments (**Fig 3**). An overview of all WPs is presented in **Fig 4**. The NORMA study is led by the Norwegian University of Life Sciences (NMBU) and conducted in collaboration with seven specialized inpatient EDs treatment units across six hospitals in Norway, located in Oslo, Bergen, Vikersund, Levanger, Bodø, and Gjøvik. Additional collaborators are The University of Oslo, Karolinska Institute and the Norwegian user organizations for eating disorders ROS and SPISFO, which has been involved in the planning of the study. WP1 includes a cross-sectional design that will investigate the microbial composition of the gut in patients with AN in comparison with HCs, and a clinical observational trial that will investigate how the composition is affected by standard care treatment in AN longitudinally (one group time-series design). We will then apply preclinical models in WP2 (in vitro fermentation experiments) and WP3 (animal experiments) based on fecal material from WP1.

**Fig 1.**
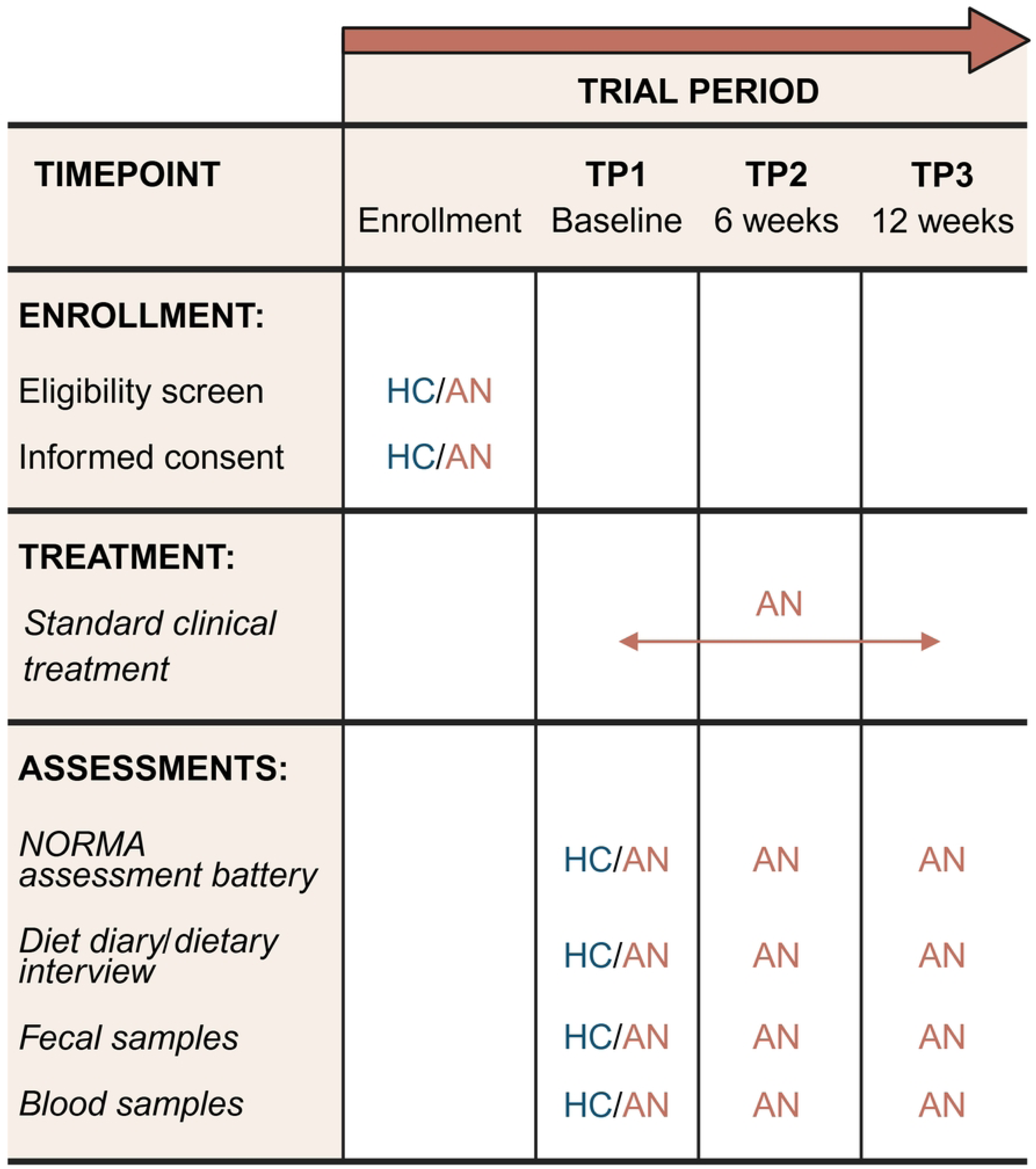
SPIRIT schedule of enrollment, treatment, and assessments in the clinical observational trial in work package 1 of the NORMA study. AN; anorexia nervosa; HC, healthy controls; NORMA, Norwegian microbiota study in anorexia nervosa; TP, timepoint. Created with BioRender.com.

**Fig 2.**
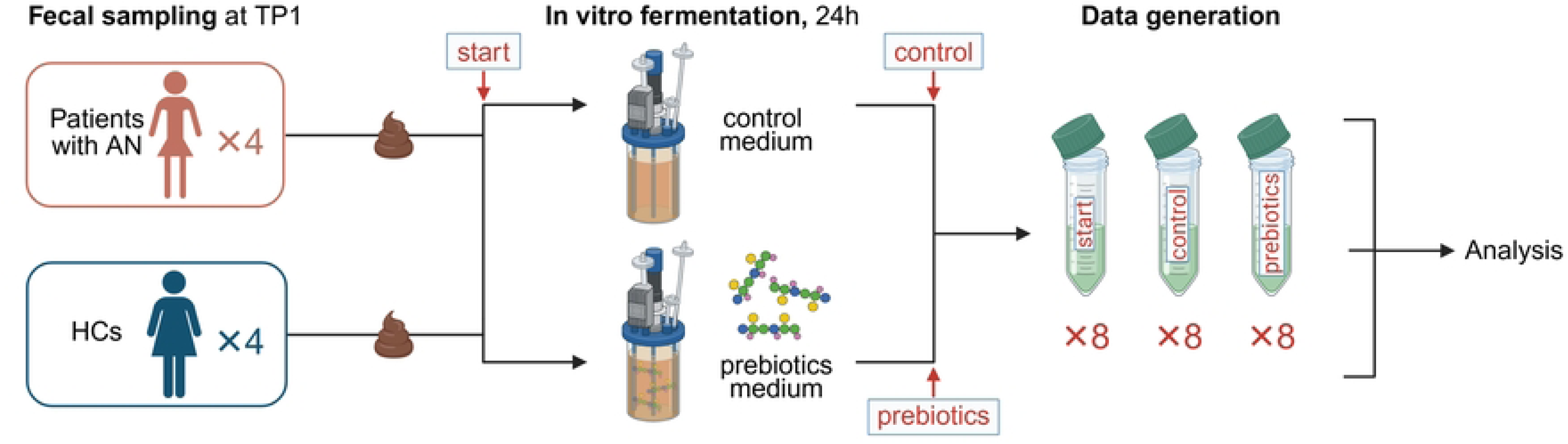
In vitro fermentation of fecal material from patients with AN and HCs in work packages 2 of the NORMA study. Samples will be anaerobically grown in a rich growth medium, either unsupplemented (control medium) or supplemented with prebiotics such as acetylated galactoglucomannan. The initial setup will include eight pre-fermentation samples (Four HCs and four patients with AN) and 16 post-fermentation samples (Four HCs and four patients with AN per growth medium). AN, anorexia nervosa; HCs, healthy controls; NORMA, Norwegian microbiota study in anorexia nervosa; TP: time point. Created with BioRender.com.

**Fig 3.**
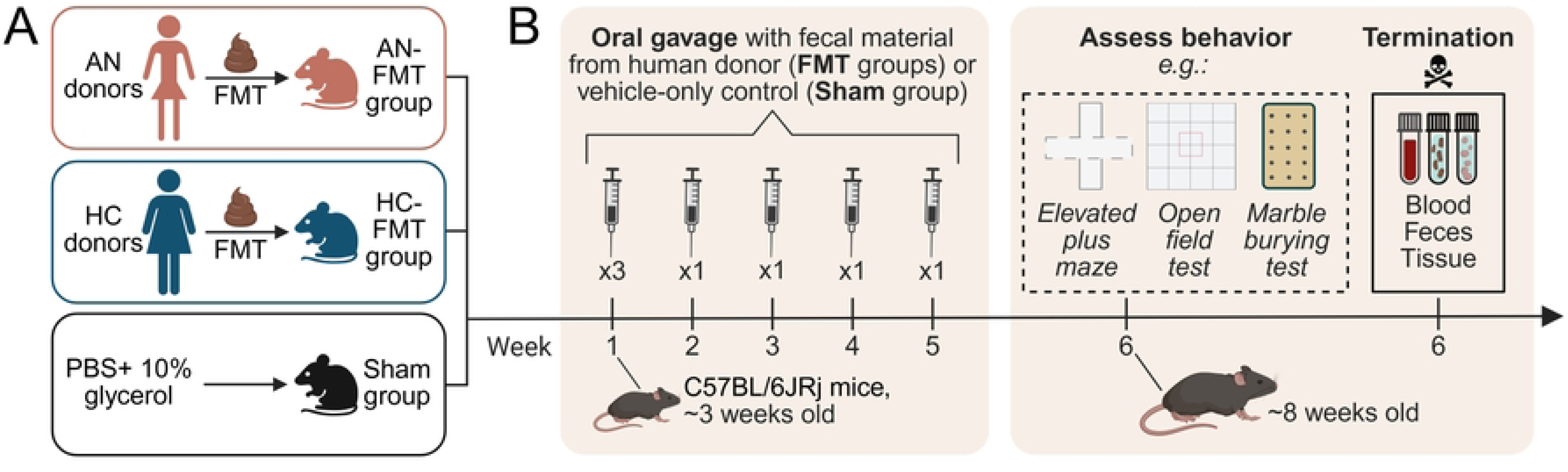
The initial animal experiment in work package 3 of the NORMA study. **(A)** Experimental groups and **(B)** Timeline. AN, anorexia nervosa; FMT, fecal microbiota transplantation; HCs, healthy controls; NORMA, Norwegian microbiota study in anorexia nervosa; PBS, phosphate-buffered saline. Created with BioRender.com

**Fig 4.**
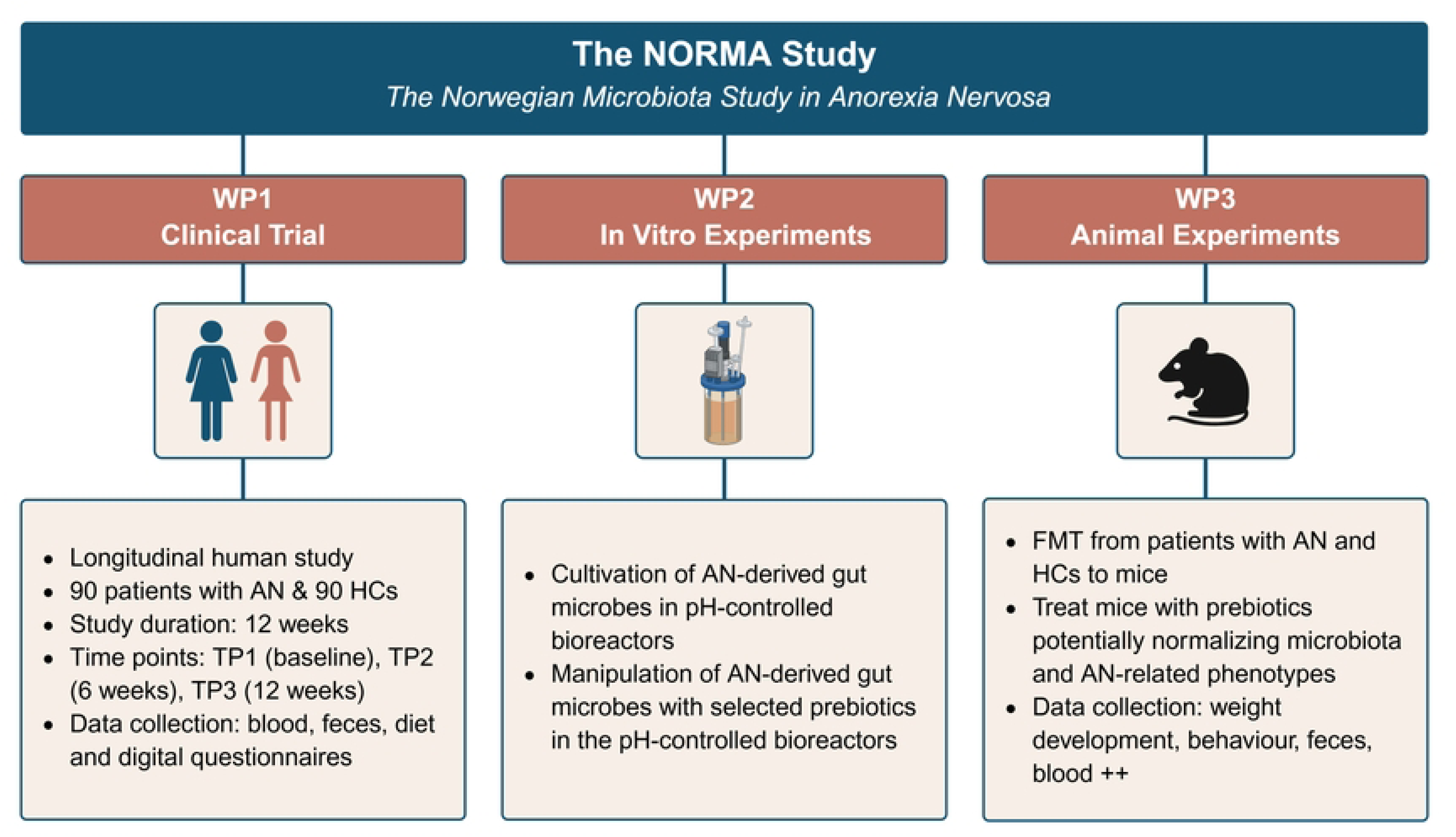
Overview of the three work packages in the NORMA study. AN, anorexia nervosa; FMT, fecal microbiota transplantation; HCs, healthy controls; NORMA, Norwegian microbiota study in anorexia nervosa; TP, time point; WP, work package. Created with BioRender.com.

Participant recruitment and data collection began on September 1st, 2023, and are expected to be completed by May 31^st^, 2026. The first results are expected to be generated during second quarter of 2026.

### The clinical trial (WP1)

#### Outcomes

Primary outcomes for WP1 are: i) differences in fecal microbiota composition between AN and HCs, and ii) changes in fecal microbiota composition-, iii) changes in mental scores-, and iv) changes in GI problems, in AN during standards care treatment at the hospital units for ED.

Secondary outcomes for WP1 are: i) associations between microbiota measures and abundance of specific taxa, serum biomarkers, dietary characteristics, GI issues and mental issues, and ii) associations between baseline microbiota composition and changes in GI complaints during standard care treatment at the clinics for ED, and iii) associations between baseline microbiota composition and changes in mental scores during the standard care treatment at the hospital units for ED.

#### Study participants

In WP1, a total of 90 patients with AN referred to specialized treatment for ED and 90 HCs will be recruited.

Patients with a primary diagnosis of AN referred to one of the six collaborating hospitals and who presumably will be under inpatient treatment for at least 12 weeks are eligible for inclusion. The collaborating hospital units include The Regional Department of Eating Disorders (Oslo University Hospital, Oslo), The Regional Department of Eating Disorders (Haukeland University Hospital, Bergen), Regional Center for Eating Disorder (Nordland Hospital Trust, Bodø), The Regional Eating Disorders Unit (Nord-Trøndelag Hospital Trust, Levanger), Modum Bad (Vikersund), District Psychiatric Center (Innlandet Hospital Trust, Gjøvik), and District Psychiatric Center (Oslo University Hospital, Søndre Oslo, Oslo). Both inpatients and patients receiving intensive day treatment are eligible for inclusion. This also includes ‘sequential inpatients’, referring to patients who are hospitalized for 4-6 weeks, then return home for a period, and are subsequently re-hospitalized. The inclusion criteria for the patients are i) female ii) diagnosed with AN (both restrictive and binge-eating/purging type), iii) age 16-50, iv) a body mass index (BMI) below 18.5 kg/m^2^, and v) sufficient proficiency in the Norwegian language to understand and complete questionnaires in Norwegian independently. The exclusion criteria are i) history of inflammatory bowel disease, celiac disease, or GI tract surgery, ii) treatment with oral antibiotics in the past two months, or iv) systematic intake of probiotic supplements in tablet form in the past two months. The inclusion and exclusion criteria are summarized in **Fig 5**.

**Fig 5.**
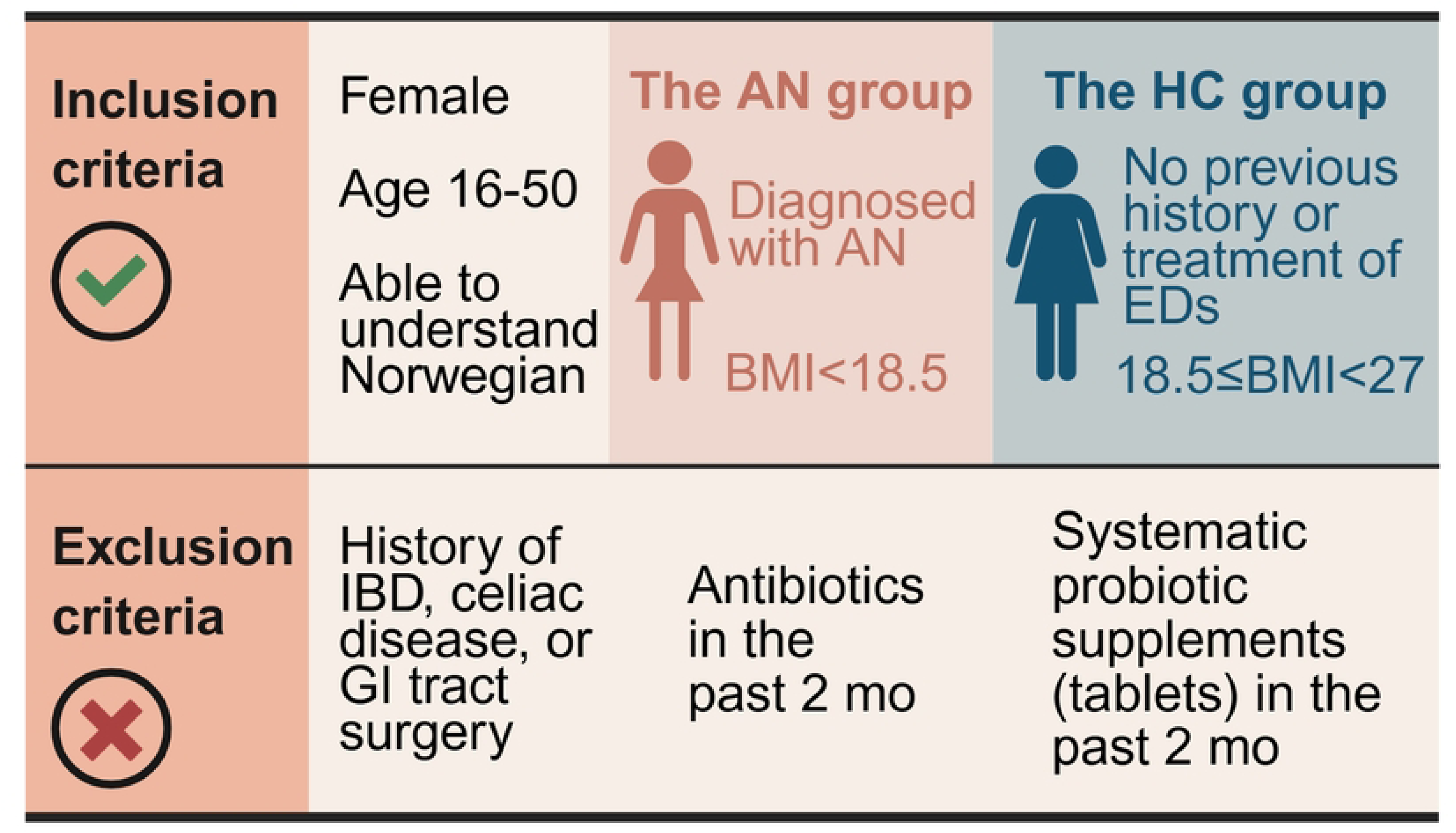
Inclusion and exclusion criteria for patients with anorexia nervosa and healthy controls in the clinical observational trial in work package 1 of the NORMA study. AN, anorexia nervosa; BMI, body mass index; ED, eating disorder; GI, gastrointestinal; HC, healthy control; IBD, inflammatory bowel disease; NORMA, Norwegian microbiota study in anorexia nervosa. Created with BioRender.com.

Recruitment of patients with AN will be conducted at each hospital unit by hospital personnel following standardized procedures and using written material provided in the study. Most patients will be informed about the study during one of their pre-admission planning meetings, typically a few weeks before hospitalization. A few days before hospitalization or, at the latest, within the first week of treatment, patients will be contacted and asked if they wish to participate. Patients who agree to participate will provide digital informed consent (**Additional file 2**), granting permission to collect biological samples and retrieve information from medical records, health registries, and questionnaires. The written informed consent will also include details about the use of individual data and the storage of biological samples during the study for analysis and publication purposes.

The HC group will include healthy individuals who meet the following inclusion criteria (Fig 5): i) female, ii) age 16-50, iii) BMI equal to or above 18.5 kg/m^2^ but less than 27 kg/m^2^ (i.e., a BMI ranging from normal weight to mild overweight), iv) no previous history or treatment of EDs, and v) have sufficient proficiency in the Norwegian language to understand and complete Norwegian-language questionnaires independently. The exclusion criteria are the same as for the patients (Fig 5).

The recruitment of 90 HCs will be conducted in two phases. An initial group of HCs will be recruited primarily via a post published on the Oslo University Hospital intranet, supplemented by strategically placed posters at key locations within the hospital. Additionally, colleagues and friends may be recruited through other social networks. The second half of the HC participants will be recruited through Ås municipality and via the intranet at NMBU. The inclusion criteria concerning age will be adapted in the second phase of recruitment to ensure a more comparable age between the HC groups and patients with AN. Potential HCs will be contacted by the study researchers by email or phone and will be screened according to the inclusion and exclusion criteria. If eligible, they will sign a digital informed consent (**Additional file 3**) and receive all necessary information for further participation in the study.

#### Overview of study design and procedures

The schedule of assessments in WP1 is presented in **Fig 1**. The included patients with AN will be assessed throughout their participation in standard clinical treatment at three time points (TP): before the start of treatment and/or within the first week of admission (TP1, baseline), and at 6– and 12-week follow-up (TP2 and TP3, respectively). Patients will be asked to provide fecal samples, fasting blood samples, and complete questionnaires about their mental and physical health, as well as record their diet at each time point. The HC participants will be assessed at one time point only (referred to as ‘TP1’). The HCs will complete approximately the same digital questionnaires as the patients, provide blood and fecal samples, and record their diet for three days. The NORMA study has approval to contact the participant within ten years after recruitment for follow-up data collection and access to medical records and registry data.

#### Data management

A data management plan has been developed for the project which has been approved by the Norwegian Research Council and was part of the approval process by the Norwegian Agency for Shared Services in Education and Research (SIKT ID number 386069). The data management plan includes information about the project, responsibilities and planning, costs and resources, data collection methods and information on data storage validation and quality control. A description of how sensitive data are handled, and ethical aspects are also included. Furthermore, the data management plan includes information on data format and software, and information about the metadata, data transfer agreements, data sharing procedures and data security. Also, routines for internal and external data storage, backup and archiving are described. Finally, the regulation of intellectual property rights is described.

#### Self-report questionnaires

Participants will receive a digital link by email to an online assessment battery consisting of eight (HC group) or ten (AN group) self-report questionnaires about different aspects of mental and physical health, GI symptoms, and stool consistency. The questionnaires are officially approved Norwegian translations. All questionnaires can be completed digitally on a computer or mobile phone. Data obtained from the online questionnaire forms will be safely collected and stored using the TSD (services for sensitive data at the University of Oslo) infrastructure and online platform. **Table 1** presents the assessment instruments, the domain captured, and the differences in the online assessment battery between the AN and HC groups.

**Table 1.** Self-report questionnaires to be used in the clinical observational trial in work package 1of the NORMA study. AN, anorexia nervosa; ED, eating disorder; HC, healthy control; GI, gastrointestinal; GSRS-IBS, gastrointestinal symptom rating scale for IBS; GAD, generalized anxiety disorder; IBS, irritable bowel syndrome; NORMA, Norwegian microbiota study in anorexia nervosa; OCI-R, obsessive-compulsive inventory revised; PHQ, patient health questionnaire; SF, short form health survey.

| Assessment | Assessed in | Domain |
| --- | --- | --- |
| NORMA background information | AN/HC | Lifetime ED, self-reported weight, use of antibiotics and probiotics, demography |
| EDE-Q6.0 | AN only | Current ED-symptoms |
| CIA.3.0 | AN only | Current psychosocial impairment |
| GSRS-IBS | AN/HC | GI complaints and IBS-symptoms |
| SF-36 | AN/HC | Quality of life |
| GAD-7 | AN/HC | Current anxiety symptoms |
| PHQ-9 | AN/HC | Current depression symptoms |
| OCI-R | AN/HC | Current obsessive-compulsive symptoms |
| NORMA Physical activity and rest | AN/HC | Physical activity and rest |
| Bristol stool scale | AN/HC | Stool consistency |

The background questionnaire (for patients with AN and HCs) includes questions that cover various aspects such as marital status, children, education level, demography, smoking and snus habits (i.e. smokeless tobacco-product), alcohol consumption, relationship with food and body image, history of antibiotic use (both lifetime and recent), medication, and use of defecation agents.

The Eating Disorder Examination Questionnaire, version 6 (EDE-Q6.0, for patients with AN only) (53) is a 28-item self-report measure adapted from the interview-based EDE and shows strong correlation with the original version (54). It assesses the range, frequency, and severity of ED-related attitudes and behaviors across four subscales over the past four weeks: restraint, eating concern, shape concern and weight concern. Higher global scores indicate more severe eating pathology, with a cut-off score above 2,5 suggestive of an ED (55).

The Clinical Impairment Assessment Questionnaire (CIA3.0, for patients with AN only) (56) is a 16-item self-report measure assessing psychosocial impairment related to ED features over the past 28 days. The questionnaire covers mood and self-perception, cognitive functioning, interpersonal functioning, and work performance. The total score ranges between 0 and 48, with higher scores indicating greater psychosocial impairment, and a global score of 16 or above considered to be clinically significant.

The GI Symptom Rating Scale for irritable bowel syndrome (GSRS-IBS, for patients with AN and HCs) (57) is a tool used to measure the severity of irritable bowel syndrome symptoms. The questionnaire consists of 13 self-report items that assess the severity of IBS symptoms across five subscales over the past seven days: abdominal pain, bloating, constipation, diarrhea, and satiety. Scores are rated on a 6-point Likert scale ranging from 1 to 7, with a total score ranging from 0 to 78. Higher scores indicate more severe IBS symptoms, categorized as minimal (0–20), moderate (21–39), and severe (40–78). Permission for use in the NORMA study has been obtained from AstraZeneca on 17th of July 2023 (**Additional file 4**).

The 36-Item Short Form Survey (SF-36, for patients with AN and HCs) (58) is a widely used self-report instrument designed to assess quality of life (QoL). The questionnaire includes 36 items that cover eight health domains over the past 4 weeks: physical activity, social activity, health problems, bodily pain, mental health, emotional problems, vitality, and general health perceptions. These are summarized into two components: physical and mental health. Higher scores reflect better QoL, though no universally accepted clinical cut-off values exist. However, scores below 40 are sometimes interpreted as indicating clinically relevant impairment.

The Generalized Anxiety Disorder Assessment (GAD-7, for patients with AN and HCs) (59) is a self-report questionnaire used to screen anxiety and assess the severity of generalized anxiety disorder. The questionnaire focuses on symptoms from the past two weeks and consists of seven questions, providing a score between 0 and 21. A score of 8 or higher is considered a reasonable cut-off for identifying probable cases of generalized anxiety disorder. The GAD-7 is a valid and reliable measure of anxiety.

The Patient Health Questionnaire 9 (PHQ-9, for patients with AN and HCs) (60) is a self-administered questionnaire that assesses symptoms of depression and is used in both clinical practice and science, and is based on the Primary Vare Evaluation of Mental Disorders (PRIME-MD) diagnostic instrument for common mental disorders. The questionnaire focuses on nine items based on the Diagnostic and Statistical Manual of Mental Disorders (DSM-IV) criteria for major depressive disorder for the last two weeks. Each symptom is scored and can give a total score between 0-27, with a score of 10 or higher as indicative of depression. The PHQ-9 is a valid and reliable measure of depression.

The Obsessive-compulsive inventory-Revised (OCI-R, for patients with AN and HCs) (61) is a self-report questionnaire consisting of 18 items which measure obsession and a variety of compulsion over the last two weeks. The symptoms are measured across six subscales; washing, checking, neutralizing, obsessing, ordering and hoarding and gives a total score between 0-72 points. A value of 21 and higher is indicative of an obsessive-compulsive disorder (OCD)-diagnosis.

The questionnaire on physical activity and rest (for patients with AN and HCs) is a self-report questionnaire consisting of eight questions about the frequency, intensity, and duration of physical activity/exercise over the past two weeks, along with 2 questions about time spent in rest/low activity. The questions on physical activity are based on a validated questionnaire from the CRC-NORDIET study (62). The purpose of the questionnaire is to assess the level of physical activity, rest, and participants’ perceptions of rest.

The Bristol Stool Form Scale (BSFS, for patients with AN and HCs) (63) is a clinical assessment tool that classifies stools into seven groups/types, which can be used as a diagnostic tool for assessing bowel health and functioning. The different types of stool consistency provide an indication of the time spent in the colon: Types 1-2 indicate constipation, Types 3-4 are considered normal, and Types 5-7 may indicate diarrhea and urgency. The BSFS chart is a valid and reliable measure of stool consistency.

#### Dietary intake

In addition to the self-report questionnaires, the patients with AN (at TP1-3) and HCs (at TP1 only) will be asked to digitally register their dietary intake for three consecutive days. The custom-made digital 3-day open food diary is based on a digital food diary developed by the University of Oslo. A link to the digital dietary assessment tool is sent to study participants via email, accompanied by oral instructions from health personnel and a video tutorial on how to complete the diet registration. The registration should be done continuously for three days, at all meals and use household measures for portion size estimation. Participants are encouraged to add pictures of their meals and to be as detailed as possible in their descriptions of all the food and drink consumed. However, the patients are asked not to take pictures at the hospital units to avoid disturbing/triggering other patients. Alternatively, patients with AN can opt for a food interview conducted by health personnel since patients with AN may find food self-registration challenging and potentially triggering. This interview is conducted as soon as possible after hospitalization and covers the last three days, or two days before hospitalization plus one day after. A standardized dietary interview guideline is used to ensure consistency across different hospital units.

The dietary intake data will be analyzed using the food, energy and nutrient database and calculation system NutriFoodCalc (NFC), at the University of Oslo. The NFC uses a comprehensive food composition database containing detailed information on the energy and nutrient content of foods commonly consumed in Norway. Each reported food item will be assigned to its corresponding entry in NFC using standardized food codes. The database will be used to compute total energy intake (KJ), the macronutrients protein, total fat (including saturated, monounsaturated, polyunsaturated), and carbohydrates (including sugars and fiber), and the micronutrients including but not limited to vitamins A, C, D, E, B-complex, calcium, iron, magnesium, zinc, and sodium. Foods will also be classified into predefined food groups according to NFCs classification system, enabling estimation of group-specific intake such as fruits and vegetables (fresh, frozen, canned), whole grains, dairy, meat and alternatives, and discretionary foods (e.g., snacks, sweets). Estimated portion sizes will be standardized and converted to grams per day using NFCs portion size references and coding guidelines. Intake will be calculated as a mean per participant per day, with nutrient values automatically computed based on both raw ingredients and cooked/processed forms, when applicable.

#### Fasting blood samples

For the patients with AN, overnight fasting blood samples will be taken at their respective hospitals, and when possible, concurrently with regular blood samples as part of normal treatment procedures. In total, six blood samples will be collected at each time point (TP1-3): three serum tubes (5.0 mL with gel), two Ethylene Diamine Tetracetic Acid (EDTA) tubes (one 4.0 mL without gel and one 5.0 mL with gel), and one 4.0 mL heparin-coated vacuum tube. All the tubes will be inverted 4-5 times immediately after blood collection, stored at room temperature (19-25°C) for minimum 30 minutes (for clotting) and maximum 60 minutes, and centrifuged (15-20 minutes, 1100-1500 ×g, 19-25°C) at the local hospital laboratories, except for the lithium heparin tube, which will be placed on ice immediately after sampling. The resulting supernatant from one of the serum samples will be aliquoted into five cryotubes at each hospital’s laboratory (i.e., five serum aliquots for each timepoint) and stored at −80°C, or for a maximum of 3 months at −20°C, before transportation to NMBU on dry ice. At NMBU, the serum aliquots will then be stored at −80°C for biobanking until further analysis. The remaining five blood samples will be analyzed at the local laboratories.

For the HCs, overnight fasting blood samples will be taken at the Regional Department of Eating Disorders at Oslo University Hospital by nurses or students who have completed an approved course in blood sampling. The HCs will collect the same blood sample types as the patients with AN, except no heparin tube, and follow the same procedures as described for the patients including aliquoting serum samples for biobanking at NMBU until further analysis. The remaining blood samples will be analyzed at the Department of Medical Biochemistry at Oslo University Hospital.

Blood serum is chosen for biobanking due to its accessibility and suitability for analyzing a wide range of biomarkers, including inflammation– and appetite-related markers. To ensure proper handling and storage of blood samples, all specimens collected at local hospitals will be processed at accredited laboratories and stored according to standard operating procedures (SOPs). Key biomarkers that will be analyzed in serum samples at the local laboratories (for patients with AN) or at the Department of Medical Biochemistry at Oslo University Hospital (for HCs) are presented in **Table 2**. Additional analyses will include, but not be limited to, biomarkers related to appetite and satiety (e.g. ghrelin, adiponectin, GLP-1, and leptin), inflammation (e.g. calprotectin, IL-1β, MCP-1, and RANTES), and gut permeability (e.g. LBP and I-FABP).

**Table 2.**
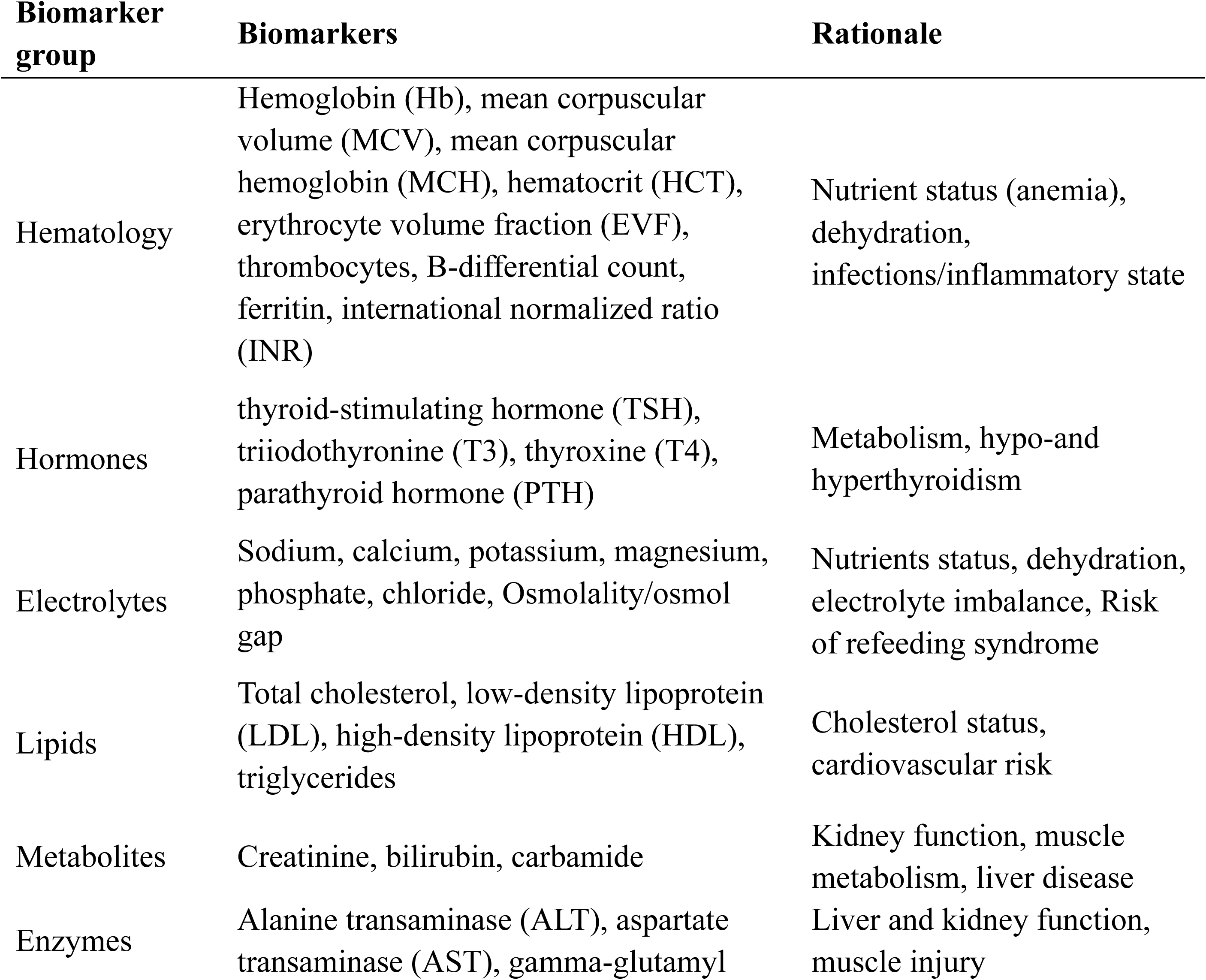
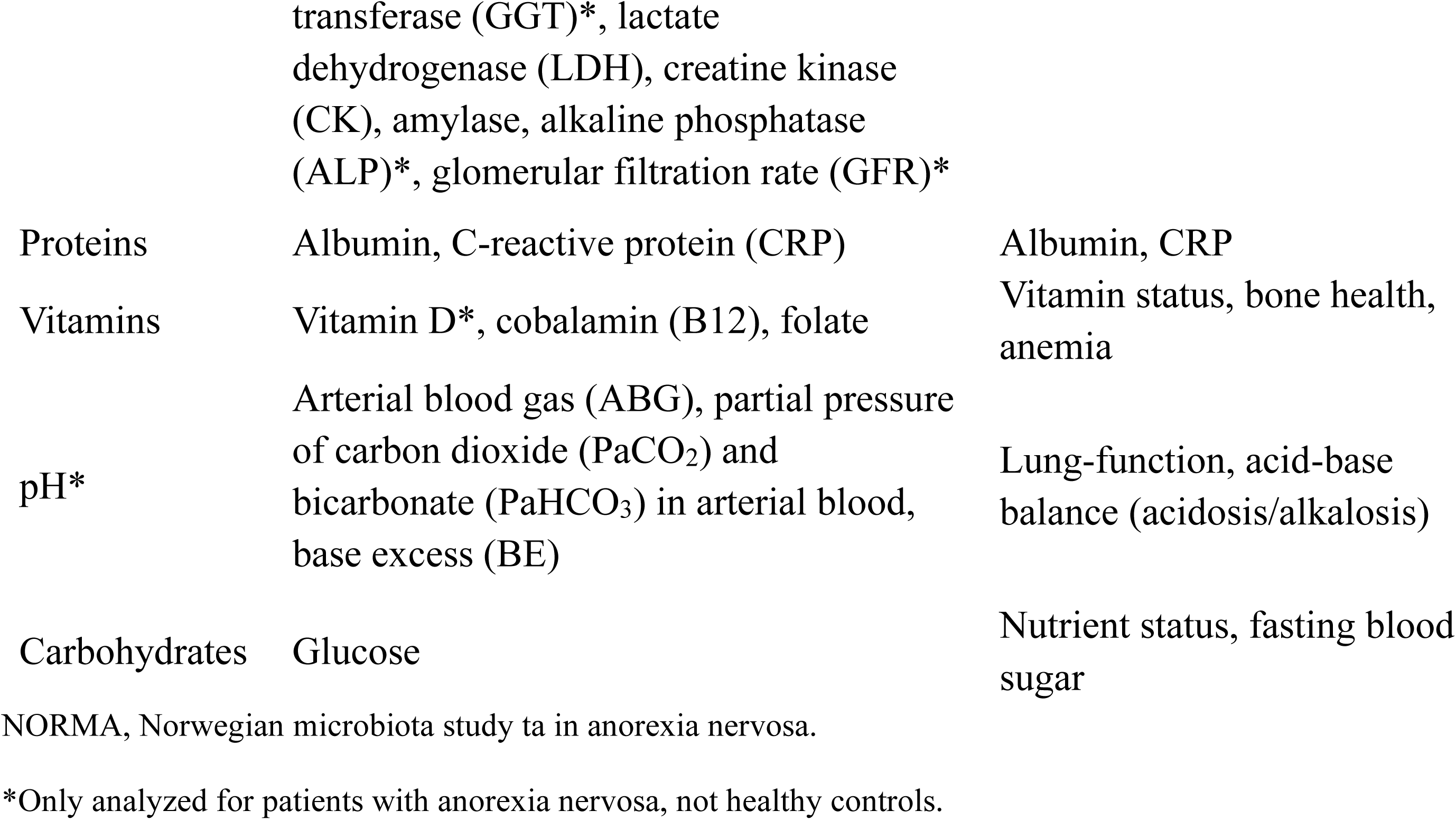
Key biomarkers to be analyzed in blood serum samples from patients with anorexia nervosa and healthy controls the clinical observational trial in work package 1 of the NORMA.

#### Fecal samples

Gut microbiota and other gut-related biomarkers will be measured using two types of fecal samples from the patients with AN and HC participants: (1) Fecal sample collected in DNA-preserving buffer (hereby referred to as ‘buffer fecal samples’) and (2) fecal samples collected in a container with an anaerobic pad (hereby referred to as ‘anaerobic fecal samples’). While buffer fecal samples will be used for biobanking and analyses such as DNA sequencing, anaerobic fecal samples will be used for in vitro fermentation experiments in WP2 and FMT mouse experiments in WP3. All patients with AN (at TP1-3) and HCs (at TP1 only) will collect two buffer fecal samples at each timepoint. Additionally, a subgroup of patients with AN from Oslo University Hospital and a subgroup of HCs will collect anaerobic fecal samples (at TP1 and TP3 for patients with AN, at TP1only for HCs). All participants will receive a fecal sample collection kit, along with a demonstration of its contents and oral instructions on proper collection procedures.

Buffer fecal samples will be collected using a Protocult Stool Collection Device (Therapak, Supplier No. 505000G) and Stool collection Tube with DNA stabilizer (INVITEK, Item No. 1038111200). Patients with AN will primarily collect buffered fecal samples upon enrollment at the clinics, preferably within one week of hospital admission (TP1), and again at the hospital at TP2 and TP3. HCs will collect fecal samples at home and send them by post to NMBU. At NMBU, the samples will be stored at −80°C for biobanking and until further processing.

Anaerobic fecal samples will be collected at the hospital unit (the Regional Department of Eating Disorders at Oslo University Hospital) for patients with AN and at home for HCs. All participants will be instructed to take a plum-sized fecal sample and place it in a container with an airtight lid. An anaerobic pad (AnaeroGen 2.5 L, Thermo Scientific) will be placed in the container to maintain anaerobic conditions. The container will then be stored in a refrigerator or cooling bag until further processing at NMBU. For the patients with AN, nurses and hospital personnel will assist with the procedure, while HCs will perform the procedure at home and temporarily store the sample in cooling bags until transportation to NMBU. Further processing of the anaerobic fecal samples will be conducted by trained students within 24 hours of sampling, following a standardized protocol at the NMBU laboratory.

The container with the anaerobic fecal sample will be opened in an anaerobic chamber (Whitley A85 Workstation; 85% N_2_, 5% CO_2_, 10% H_2_) or home-made anaerobic box (dry ice at the bottom and continuous flushing with CO_2_ and N_2_), and two 4-gram portions of fecal material will be weighed out. For WP3, 4 g fecal material will be dissolved in 40 mL freshly made reduced PBS (i.e., 45 mL anaerobic Dulbecco’s phosphate buffered saline (Sigma-Aldrich, #D8537), mixed with 45 mg L-cysteine hydrochloride (Sigma-Aldrich, #C1276), and 5 mL glycerol (Sigma-Aldrich, #G5516)), whereas the 4 g samples destined for WP2 will be dissolved in an anaerobic cryoprotective solution (0.1 M phosphate buffer pH 6.8, 15% (v/v) glycerol, 5% (w/v) sucrose, 0.1% (w/v) L-cysteine hydrochloride, 0.03% (w/v) riboflavin). Both suspensions will be homogenized at room temperature for five minutes at 30 Hz using Mixer Mill (MM 400, RETSCH). Next, the homogenates will be filtered (100 μm pore size), aliquoted (20 aliquots á 1.5 mL for WP2, 30 aliquots á 1 mL for WP3), snap frozen in liquid nitrogen, and stored at −80°C until use in the fermentation (WP2) and animal (WP3) experiments. If there is residual material, an aliquot of at least 0.6 g from the anaerobic fecal sample will be collected for SCFA analysis. Any excess fecal material from the anaerobic fecal sample beyond this will be stored in the NMBU biobank at −80°C. The protocol for the collection and processing of the anaerobic fecal samples was developed based on Bokoliya et al. (64) with the goal to ensure that the microbes in the samples will not be exposed to significant amounts of oxygen during processing.

#### Anthropometric measurements and medical records

For the patients with AN, anthropometric measurements (body height and weight, and body weight history) will be gathered from medical records at each hospital. Results from blood serum analyses and information about medications, including the use of laxatives and dietary supplements, will also be recorded from the patient’s medical records. HCs will report their body weight and height as part of the self-report questionnaires but will be offered the opportunity to have new measurements taken at the hospital if necessary. Anthropometric measurements should be taken within a week of each time point.

#### National health registries

The NORMA study has the approval to collect information about the participants’ through the Norwegian Prescription Database, the Norspis registry (Norwegian Quality Register for the Treatment of Eating Disorders), the Norwegian Patient Register, and the Norwegian Registry for Primary Health Care – used to record contacts with health services related to mental health treatment) at TP1-3 and after 1, 5 and 10 years after enrollment in the study. These data can provide detailed insights into antibiotic use and other relevant medication use for both the HC group and AN group, pertinent to the NORMA study.

#### Compliance strategies

Patients with AN will receive a gift card worth 200 NOK for each completed time point. They will be informed about the gift cards at the time of recruitment and will be made aware that participation will involve additional follow-up and contact with study personnel during the study period. After participating in the study, the HCs will receive a personalized health report including information about their blood tests and nutritional status. Additionally, five gift cards worth 1000 NOK each will be given to randomly selected HCs at the end of the study.

#### Sample size calculations

The sample size needed to detect statistically significant differences in the fecal microbiota composition between patients with AN and HCs at TP1 was estimated using a web-interface for simulation-based power calculations. A Dirichlet-Multinomial model is used to describe and generate abundances (fedematt.shinyapps.io/shinyMB) using the default setting except # of operational taxonomic units (OTUs) was set to 100, significance level (α) was set to 0.05, and the abundance curves were made based on the five most abundant OTUs increased by 60% and the five next most abundant OTUs increased by 50%. With these settings, the sample size required to detect a statistically significant difference between the patients with AN and HCs with 80% power will be 79 participants in each group. With an estimated drop-out of ∼10%, we plan to include 90 patients with AN and 90 HCs.

#### Hypotheses and planned statistical data analyses in WP1

The clinical observational trial in WP1 is designed to test the following hypotheses.

WP1, Hypothesis 1: The gut microbiota in patients with AN is different from that of HCs. We will compare different indices of alpha diversity, investigating both microbial richness and evenness, and different measures of beta diversity. Also, differences in bacterial abundances at various taxonomic levels (e.g. phylum, class, order, family, and genus) will be investigated using conventional statistical approaches as well as more advanced multivariate methods.

WP1, Hypothesis 2: Inpatient treatment of patients with AN combining cognitive treatment with re-nutrition strategies will change the gut microbiota in a more normalized direction (more similar to HCs). Mixed models will be applied to statistically analyze the effect of time on different indices of alpha diversity and bacterial abundances at various taxonomic levels (e.g. phylum, class, order, family, and genus) with and without adjusting for baseline characteristics such as age and BMI. Differences in the overall bacterial communities between the patients with AN at the different visits and HCs based on the beta diversity measures will be assessed using methods such as global permutation based multivariate analysis of variance (PERMANOVA).

WP1, Hypothesis 3: Gut microbiota composition in patients with AN at TP1 (before treatment) is associated with serum biomarkers such as biomarkers of appetite regulation and inflammation, dietary characteristics, GI issues, and mental issues, and can be used to predict treatment success in terms of improved BMI and EDE-Q-scores. Associations between relevant parameters will be analyzed using multivariate analysis such as principal components analysis (PCA), canonical variate analysis, independent components analysis, and multivariate regression. Regression models will be used to investigate associations between microbial diversity and clinical outcomes, adjusting for age and BMI. Machine learning approaches such as partial least squares regression, gradient boosting, or artificial neural network algorithms will be used to predict the response variables (clinical outcomes and treatment success), from the large set of predictor variables; microbial taxa, gene, or pathway content.

WP1, Hypothesis 4: Changes in microbiota composition during inpatient treatment towards that of HCs are associated with improvements in BMI as well as improvements in dysregulated serum biomarkers, GI complaints, and mental health. Associations will be analyzed using similar strategies as described in hypothesis 3.

### The in vitro experiments (WP2)

#### Experimental groups

To test the ability of the fecal microbes to ferment prebiotics, processed anaerobic fecal samples from patients with AN and HCs, collected in WP1 (see section “Fecal samples”) will be cultured in vitro as part of WP2. We will use acetylated galactoglucomannan as the pioneer prebiotic supplement and test impact on the microbial communities using fecal samples from four patients with AN and four HCs (**Fig 2**).

#### Bioreactors

The bioreactors to be used in the WP2 experiments are produced in-house and consist of a double-jacketed stainless-steel vessel with a lid made of polyether ether ketone (PEEK), containing connection ports for gas, acid and base, and a gas-sealed septum sampling. The bioreactors will be pH-controlled (6.8±0.2 by automatic addition of 1 M HCl and 1 M NaOH), the headspace will be continuously flushed with N_2_ gas (O_2_ < 5 ppm), the temperature will be maintained at 37°C via a circulation bath (Thermo Fisher Scientific Inc.), and the content will be stirred at 300 rpm.

#### Preparations

Sterile basal fermentation media containing prebiotics (0.5 % w/v) or ddH_2_O as control will be made according to **Table 3**, adjusted to pH 6.8, and 70 mL will be transferred to each of the autoclaved bioreactors in a sterile laminar flow hood. The bioreactors with media, alongside HCl and NaOH, will then be placed in an anaerobic chamber (Whitley A85 Workstation; 85% N_2_, 5% CO_2_, 10% H_2_, 37°C) for at least 12 hours to become anaerobic. Sterile pH probes will be inserted into the bioreactors before inoculation. The processed anaerobic fecal samples from WP1 will be thawed in the anaerobic chamber, and optical density (OD_600_) will be measured and used to adjust inoculation volume.

**Table 3.** Concentration of the different components of the sterile basal fermentation media that will be used for in vitro fermentation experiments in work package 2 of the NORMA study.

| Component | Concentration |
| --- | --- |
| Peptone water | 2 g/L |
| Yeast extract | 2 g/L |
| NaCl | 0.1 g/L |
| K<sub>2</sub>HPO<sub>4</sub> | 0.04 g/L |
| KH<sub>2</sub>PO<sub>4</sub> | 0.04 g/L |
| MgSO<sub>4</sub>*7H<sub>2</sub>O | 0.01 g/L |
| CaCl<sub>2</sub>*6H<sub>2</sub>O | 0.01 g/L |
| NaHCO<sub>3</sub> | 2 g/L |
| Hemin | 0.05 g/L |
| Vitamin K | 10 µ/L |
| L-cysteine HCl | 0.5 g/L |
| Sodium glycocholate | 0.5 g/L |
| Sodium taurocholate | 0.5 g/L |
| Prebiotic/carbohydrate source | 0.5 % w/w |
| ddH<sub>2</sub>O | To 1 L |

NORMA, Norwegian microbiota study in anorexia nervosa

#### Fermentation

Based on the OD_600_ measured during the preparation, the processed anaerobic fecal samples from WP1 will be inoculated to a final OD_600_ of 0.4 into the bioreactors while still in the anaerobic chamber. After 1 hour, the bioreactors will be transferred out of the anaerobic chamber, placed on magnetic stirrers, and connected to the gas inlets and outlets, pH-adjustment tubes, and the water bath. The fermentation will run for 24 hours. Samples taken during the fermentation will be mixed (1:1) with RNAlater (Sigma-Aldrich).

#### Hypotheses in WP2

The in vitro fermentation experiments in WP2 will be designed to test the following hypothesis.

WP2, Hypothesis: Carbohydrates with prebiotic potential (e.g., galactoglucomannan) shown to stimulate the growth of bacterial species that have low abundance in AN (e.g., *Roseburia intestinalis, Faecalibacterium prausnitzii, Blautia spp, Bifidobacterium spp, Agathobacter faecis* also known as *Roseburia faecis*) will increase the abundance of these beneficial bacteria in the in vitro culture of AN-derived microbiota.

### The animal experiments (WP3)

#### Mouse groups and timeline

In WP3, anaerobic fecal samples from patients with AN and HCs, collected in WP1 (see section “Fecal samples”), will be used as donor material for FMT into newly weaned female C57BL/6JRj mice. An initial experiment will include three groups of mice (**Fig 3A**): AN-FMT, HC-FMT, and Sham. Mice in the AN-FMT and HC-FMT groups will receive FMT from AN and HC donors, respectively, via oral gavage. Fecal material from eight AN and eight HC donors will be used for this purpose, and four mice will receive FMT from each donor. All mice in the Sham group will, instead of FMT with fecal material, receive a solution of phosphate-buffered saline with 10% glycerol, serving as a vehicle-only FMT control. Apart from this, the oral gavage protocol will be the same for all mice across the three experimental groups.

The timeline for the initial mouse experiment can be seen in **Fig 3B**. Based on the results from an FMT experiment reported by Le Roy et al. (65), the mice will be approximately three weeks old (i.e., at weaning) at the start of the experiment, targeting the critical period in gut microbiota establishment known as ‘the window of opportunity’. During the first experimental week, mice will receive FMT or Sham treatment on three consecutive days, as done in Le Roy et al. (65). In the next four weeks, FMT or Sham treatment will be administered once weekly, in accordance with another experiment with FMT from AN donors to mice (38). After the five-week treatment period, the mice will undergo behavioral tests to assess anxiety– and compulsive-like (i.e. repetitive) behaviors using tests such as the open field (OF), elevated plus maze (EPM), and marble burying (MB) tests. Body weight and food consumption will be recorded weekly during the experiment, and fecal droppings will be collected before the first FMT/Sham treatment and at the termination day. After euthanasia, blood, intestinal content, brain tissue, and other types of relevant tissues will be collected.

Additional mouse experiments will be performed based on the results of the initial mouse experiment in WP3 and the findings in WP2. Ideally, follow-up experiments will aim to determine whether gut microbiota-modulating interventions, such as prebiotics used in WP2, can normalize AN-associated alterations in the gut microbiota of AN-FMT mice. Such normalization may, in turn, influence other AN-related traits, including behavioral and metabolic phenotypes. However, these experiments will only be pursued if the FMT in the initial experiment successfully induces both AN-associated microbial profiles and relevant phenotypic traits in the recipient mice.

#### Fecal microbiota transplantation

Oral gavage with fecal material from patients with AN or HCs (in the AN-FMT and HC-FMT groups, respectively), or with PBS containing 10% glycerol (in the Sham group), will be performed a total of seven times during the initial mouse experiment (Fig 3B). The processed anaerobic fecal samples from WP1 (see section “Fecal samples”) will be thawed at approximately 37°C and thoroughly mixed. Thawed samples will be used for oral gavage within 2 hours, using a 24G ball-tip steel gavage needle. Each oral gavage administration will consist of 100 µL, an appropriate volume for mice weighing 10 g or more.

#### Assessing anxiety– and compulsive-like behaviors in mice

In mice, anxiety– and compulsive-like behaviors can be assessed using several well-established behavioral tests, including the OF, EPM, and MB tests (66) (**Fig 6**). The EPM consists of a plus-shaped apparatus elevated above ground, with two open arms and two closed arms. The EPM is a well-established behavioral test that assesses anxiety-like behavior based on the natural conflict in rodents between the drive to explore and the aversion to bright, open, and elevated environments. The open arms, combined with the elevated platform, create a mildly aversive environment that enables assessment of this exploration-avoidance conflict. More time spent in the open arms is interpreted as reduced anxiety-like behavior, whereas preference for the closed arms indicates higher anxiety levels. The OF consists of a square box with four walls and assesses anxiety-like behavior and locomotor activity, based on the assumption that anxiety suppresses an animal’s drive to explore novel environments, while reduced anxiety promotes exploration. One commonly observed behavior in the OF test is thigmotaxis, meaning the tendency to remain close to the walls and corners of the arena while avoiding the exposed center. Mice that exhibit higher locomotor activity, increased rearing, and reduced thigmotaxis are generally interpreted as less anxious and more explorative. The MB test is conducted in a standard bedding-filled cage, where a set number of glass marbles are evenly spaced on the surface of the bedding. The MB test primarily assesses compulsive-like, repetitive behavior in rodents, and to some extent, anxiety-related responses. The tendency to bury marbles is considered a form of species-typical digging behavior that may be exaggerated in the context of heightened arousal or compulsivity.

**Fig 6.**
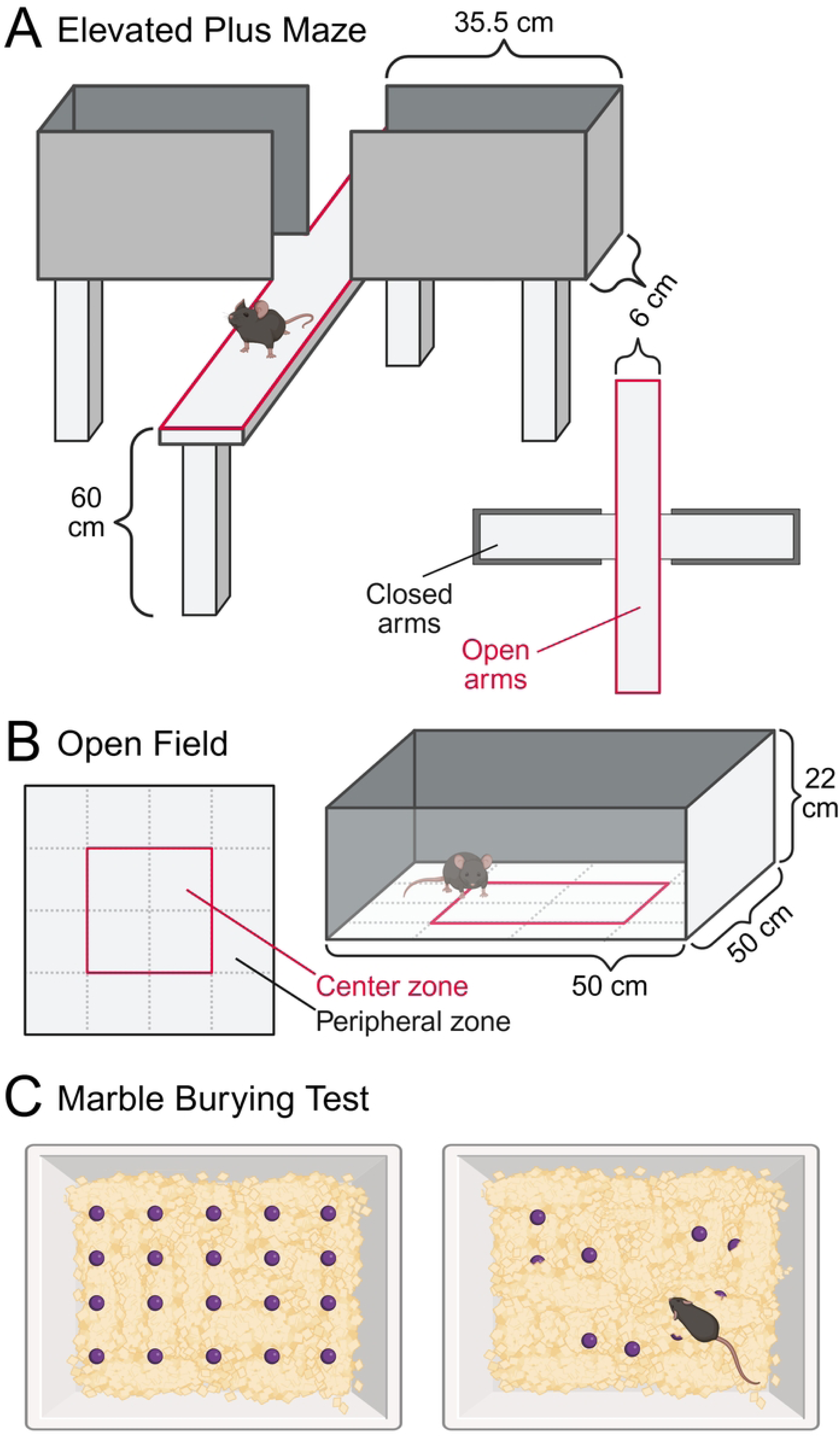
Behavior tests for assessing anxiety– and compulsive-like behaviors in mice. **(A)** Elevated plus maze **(B)** Open field test **(C)** Marble burying test. Created with BioRender.com.

#### Sample size estimation: number of FMT donors and recipient mice

The initial experiment in WP3 will have 32 mice in the AN-HC group (eight AN donors, four mice per donor), 32 mice in the HC-FMT group (eight HC donors, four mice per donor), and approximately 6 mice in the Sham group. Thus, within the AN-FMT and HC-FMT group, there will be eight “donor groups” with four mice in each. The number of mice in the AN-FMT and HC-FMT groups was determined through power analyses based on previously published studies with similar research objectives (67, 68), together with our own experience (not published), to ensure adequate statistical power. The OF behavioral variable total distance (cm) moved was used as response variable in the power analyses. It was assumed that the total distance moved will be on average 34% lower in the AN-FMT than in the HC-FMT group. Additional details regarding the sample size calculations for WP3 are provided in the Supplementary Method Description (**Additional file 5**).

#### Hypotheses in WP3

The mouse experiments in WP3 are and will be designed to test the following hypotheses.

WP3, Hypothesis 1: Mice receiving gut microbiota from patients with AN via FMT (AN-FMT mice) will exhibit different responses compared to mice receiving gut microbiota from HC donors (HC-FMT mice). These responses (hypothesis) are expected to include but not be limited to food intake (lower in AN-FMT vs. HC-FMT mice), body weight development (less increase in AN-FMT vs. HC-FMT mice), anxiety– and compulsive-like behaviors (more in AN-FMT vs. HC-FMT mice), alterations in gene expression profiles in discrete brain regions connected to anxiety, and appetite and metabolic regulation, and gut-barrier functions (impaired in AN-FMT vs. HC-FMT mice).

WP3, Hypothesis 2: AN-related phenotypes in AN-FMT mice, such as reduced appetite, impaired body weight development, elevated anxiety– and compulsive-like behavior, and disrupted gut health, can be reversed by modulating the gut microbiota through targeted prebiotic treatment.

## Discussion

By joining forces of researchers, clinical health care services, and voluntary sector, the NORMA study aims to improve our understanding of the role of the gut microbiota in patients with AN. Together, the three WPs that comprise the NORMA study—a clinical observational trial (WP1), in vitro fermentation experiments (WP2), and animal experiments (WP3)—will offer a unique, multidimensional, and transdisciplinary approach to investigate the gut microbiota and its interaction with GBA in AN. Not only will we consider factors like diet, medications, and GI complaints (WP1), but we will also, in the preclinical experiments (WP2 and WP3), explore potential underlying mechanisms and therapeutic targets, such as prebiotic treatments. The results of the NORMA study will therefore pave the way for more effective and tolerable weight restoration strategies, as a long-term goal. To our knowledge, the targeted use of prebiotics to normalize AN-related microbiota dysbiosis has not been explored previously.

While the fecal material collected in WP1 is the foundation for the preclinical experiments in WP2 and WP3, the main aim of the clinical observational trial is to explore the gut microbiota in patients with AN before, during, and after standard care treatment. We consider these investigations crucial due to the relatively few studies so far that have examined the relationship between gut microbiota and AN (19, 33, 48). Importantly, many of the previous studies of gut microbiota in AN have significant limitations, including small sample sizes (9 out of 15 studies with n≤25), reliance on cross-sectional designs (18, 27, 30, 31, 69, 70), and the absence of control groups (34, 71, 72). Furthermore, among the largest studies, which include between 55-93 patients with AN (11, 18, 24, 26, 32, 71), essential factors like diet (18, 24, 32), antibiotic use (24), and other variables that significantly influence the gut microbiota composition are often inadequately addressed, as highlighted in two recent systematic reviews (19, 33). The use of different methodologies and the absence of a standardized definition of ‘gut microbiota dysbiosis’ further complicate interpretations (33) and make it difficult to identify therapeutic targets to develop microbiota manipulative treatments.

Due to the limitations of the previous studies on gut microbiota in patients with AN, there is a clear need for larger-scale, longitudinal studies. While existing research suggests that microbial mechanisms may play a role in the etiology of AN (18, 26), more evidence is needed to determine whether gut microbiota dysbiosis is a consequence of the disorder, a contributing factor to its onset, or involved in its maintenance. The NORMA study will address these gaps by combining a well-powered clinical observational trial accompanied by animal and in vitro experiments, integrating detailed dietary and medical data, and standardized and validated methodologies. This approach allows for a more accurate assessment of microbiota changes over time but also provides insights into the underlying mechanisms. The remainder of the discussion outlines the core strengths of the NORMA study, as well as key methodological aspects to consider in relation to the current research protocol.

In addition to being one of the largest longitudinal studies of gut microbiota in AN—including patients from six hospitals across Norway— the NORMA clinical trial benefits from several major methodological strength: firstly, the high adherence of all patients—primarily inpatients—to specialized treatment programs for EDs throughout the research period. This adherence results in a consistent treatment framework, including structured routines, and dietary interventions across the patient cohort. Although minor differences in therapeutic approaches and nutritional treatment will exist across hospitals, the overall consistency will reduce the risk of confounding factors. This uniformity represents a significant methodological advantage compared to studies primarily involving outpatient populations, where treatment heterogeneity is typically greater.

Furthermore, the data collection in WP1 is conducted at three time points, allowing us to distinguish between the acute effects of re-nutrition after 6 weeks and the more long-term effects after 12 weeks of treatment. Some studies describe a ‘microbiota shift’ during treatment (25, 26, 32), but more insight is needed to pinpoint when this shift occurs, as only one of the five longitudinal studies in the field includes more than two time points for data collection (26). It should also be noted that the NORMA study has received approval to track patients for up to ten years after enrollment. This includes permission to collect additional data from medical registries and hospital readmissions, providing a valuable opportunity for long-term analysis. This will provide the opportunity to further explore whether the gut microbiota could serve as a prognostic marker for readmission, recovery, and other AN-relevant outcomes, as proposed by Andreani et al. (26).

The NORMA clinical trial also stands out from previous studies by thoroughly assessing antibiotic use in childhood and prior to treatment, and by carefully defining exclusion criteria for antibiotic use (i.e. treatment with oral antibiotics in the past two months). While some studies set this exclusion period at four weeks or less (25, 26, 30), or not specify previous use of antibiotics (24, 31), the NORMA study has opted for a longer timeframe of two months to further reduce the possibility that antibiotics influence the results.

Additionally, the inclusion of a control group at baseline is crucial due to the lack of a ‘gold standard’ for healthy gut microbiota and inconsistent definitions of dysbiosis (33). This baseline comparison helps identify problematic aspects of AN gut microbiota. Since the core gut microbiota in healthy individuals is relatively stable over time (73), HCs are sampled at one time-point only. HCs will mainly be recruited from healthcare workers and students in south-eastern Norway. As most healthcare workers and students have moved from other regions, this will ensure a broad geographic diversity and minimizing potential regional bias in gut microbiota composition.

Finally, a key strength of the NORMA clinical observational trial is the comprehensive and detailed dietary registration, which is often lacking in previous studies. This omission in the literature is paradoxical, given how central diet is in modulation of the gut microbiota (20, 74) and the fact that changes in eating behaviors and diet, are key characteristics of AN (22). By carefully documenting dietary intake, the NORMA study may determine the extent to which dietary factors explain the dysbiosis observed in AN. The combination of using an open and detailed dietary assessment method and conducting dietary interviews performed by trained personnel or dietitians ensures the quality and reliability of dietary data. Furthermore, the study includes systematic mapping of medication use, allowing for assessment of potential associations between medication and microbiota dysbiosis, an area that, to our knowledge, has not yet been thoroughly explored in this population (19).

While the NORMA observational clinical trial will address many of the shortcomings of previous studies, some important limitations and methodological considerations must be addressed. Firstly, the study duration is limited to 12 weeks of standard care treatment which may restrict conclusions about long-term outcomes and the extent to which underweight is an explanatory factor of microbiota dysbiosis. Secondly while the majority of the included patients will be inpatients, the patient population will also include some individuals receiving sequential treatments. However, while this approach may lead to less control over the treatment framework, it could also result in greater variability and enhance the generalizability of the findings to a broader range of patients with AN. Thirdly, a clear scope restriction is that the NORMA study focuses exclusively on females with severe AN. This decision is based on the higher prevalence of AN among females, who are disproportionately affected by its most severe forms. Additionally, including only severely ill patients allows for the advantage of studying participants in the controlled environment of specialized treatment wards, ensuring standardized care and minimizing external variables. However, this focus may reduce the generalizability of the findings to males, individuals with other EDs, and patients with less severe forms of AN, as the study population may not fully represent the broader AN patient group.

To uphold ethical research standards, the NORMA study employs in vitro experiments and in vivo ‘humanized’ mouse models in place of direct dietary interventions in patients with AN. This approach minimizes patient burden by avoiding disruptions to standard clinical treatment and reducing stress and anxiety. Furthermore, it allows for the investigation of microbiota interactions in a controlled environment, circumventing the ethical complexities associated with experimental dietary manipulation in this vulnerable population The in vitro experiments using human fecal microbiota allow for the identification of potential therapeutic targets and prebiotic treatments that can be used to manipulate the microbiota community. The prebiotic galactoglucomannan, chosen for this study, efficiently boosts the growth of butyrate-producing bacteria like *Roseburia intestinalis* and *Faecalibacterium prausnitzii*, which may be considered key targets for microbiota improvements (52) and is an important first step in exploring whether microbiota from AN would benefit from such exposure.

However, the clinical relevance of the in vitro experiments will be limited by several factors. Notably, cultivating gut microbes in bioreactors also presents challenges in replicating key physiological conditions, such as dynamic flow, microbial turnover, and spatial gradients in oxygen and pH. A particular concern is the potential loss of strict anaerobic bacteria and rare taxa, as culture-based methods can fail to preserve the full microbial diversity found in the original sample. To mitigate this, the NORMA study will adapt a rigorous protocol for sample collection and biobank preparation. Fecal samples from WP1 will be maintained under anaerobe conditions from the moment of collection until they are transferred into the anaerobe chamber for use in WP2 and WP3. Nevertheless, the culture-media employed may still induce ‘artificial’ shifts in the microbial community composition, favoring the growth of certain microbial taxa over others. Fast-growing species are likely to be overrepresented, although this remains to be systematically evaluated.

Additionally, the artificial and simplified nutrient input in the in vitro systems can only mimic aspects of local microbial metabolism and lack the complexity of the in vivo GI environment. Particularly, the in vitro system will not account for metabolic and immunological interaction with the host, such as via the GBA. The NORMA animal experiments will thus ensure that causalities are tested in a more relevant biological context which will be more representative of a human response. Still, although animal experiments and set ups for exploring behavior have been rigorously tested and utilized in numerous studies, a direct transfer to humans must be interpreted with caution. Both OF and EPM behavior tests have revealed strengths and limitations in assessing anxiety like behaviors, especially in the context of testing drugs aimed at reducing anxiety (75), which we have considered carefully in the current experimental design. Including the MB test as a measure of compulsive-like repetitive behaviors must also be considered with caution. However, employing a battery of mouse behavioral tests with complementary qualities will most likely generate a better overall picture and sensitivity for assessing potential behavioral differences imposed by microbiota differences between patients with AN and HCs.

Taken together, while the applicability of these preclinical models to humans remains uncertain, they serve as an important first step in identifying promising prebiotic supplements. If positive effects of prebiotic candidates are observed on the cultivated microbiota, future clinical trials may further explore these prebiotics in patients with AN through carefully designed supplement-based or dietary interventions.

In summary, investigating the relationship between gut microbiota, core symptoms, and dietary factors in AN is crucial, as identifying microbial patterns and their response to treatment could make gut microbiota modulation a valuable component of AN management. The NORMA study will be among the largest investigations into gut microbiota changes in response to AN treatment and uniquely integrates clinical and preclinical research to explore the role of gut microbiota in AN. By expanding knowledge in this field, there is potential to pave the way for innovative treatments, such as prebiotics and tailored nutritional guidelines, that could be seamlessly integrated into clinical practice. By addressing key factors like diet and use of medication, the study aims to generate essential insights that could lead to more effective treatments and improved patient outcomes.

## Declarations

### Ethics approval and consent to participate

This study was conducted according to the guidelines laid down in the Declaration of Helsinki, and all procedures involving human participants (in WP1), and participant-derived samples and data (in WP1-3), were approved by the Regional Committee for Medical and Health Research Ethics, South East A on June 27, 2023 (REK ID number 588768) and Norwegian Agency for Shared Services in Education and Research (SIKT ID number 386069). The NORMA study is also registered with the National Institutes of Health Clinical Trials (www.ClinicalTrials.gov; Identifier: NCT06144905, registration date: September 22, 2023). All participants, both patients with AN and HCs, provided informed online consent to participate. Unless consent is actively subtracted, all data and material collected can be used in the NORMA study, including follow-up information collected from the National health registries. The initial mouse experiment in WP3 will be performed with permission from The Norwegian Food Safety Authority (FOTS #31150), and will be conducted in compliance with the current guidelines of The Federation of European Laboratory Animal Science Associations. Protocol modifications involving WP1 and WP3 must be approved by REK and FOTS respectively. Authorships will follow Vancouver guidelines.

### Consent for publication

All participants, both patients with AN and HCs, provided informed online consent for publication of the results.

### Availability of data and materials

This manuscript does not report data generation or analysis.

The datasets generated and analyzed during the NORMA study will not be publicly available due to ethical and legal considerations. Specifically, participants did not provide written consent for their data to be shared in open-access repositories or with third parties beyond the scope of the approved research protocol.

However, de-identified data and relevant study materials may be made available by the corresponding author upon reasonable request, subject to appropriate institutional approvals and in compliance with applicable data protection regulations (e.g., GDPR). Any data sharing will require a data use agreement that ensures the protection of participant confidentiality and respects the ethical obligations under which the data were collected.

### Plan for dissemination

Results from the project will first be disseminated through traditional scientific channels including high-impact journals and conferences. We will focus on sharing research results with the users: patients, patient organizations, health personnel, peers in the research field, policymakers and the general public. After the completion of recruitment, **t**he patients in the study will receive an annual update from the study. The two user organizations that represent the societal challenge of the project will assist in the translation of the scientific papers into popular science and help identify the most efficient publishing platforms for result communication to the public.

### Authors’ contributions

IHH (Conceptualization, data curation, investigation, methodology, project administration, resources, visualization, writing-original draft)

LB (Conceptualization, funding acquisition, supervision, writing-review and editing)

AMH (Data curation, methodology, project administration, visualization, writing-original draft)

SSS (Methodology, supervision and writing-review and editing)

TBS (Resources, writing-review and editing)

AJ (Resources, writing-review and editing)

KLO (Resources, writing-review and editing)

DA (Resources, writing-review and editing)

TEA (Resources, project administration, writing-review and editing)

YMV (Methodology, writing-review and editing)

JHMS (Writing-review and editing)

MHC (Methodology, writing-review and editing)

MLO (Resources, writing-review and editing)

RA (Resources, writing-review and editing)

KH (Project administration, resources, writing-review and editing)

SW (Project administration, writing-review and editing)

IS (Project administration, writing-review and editing)

JD (Project administration, writing-review and editing)

HTR (Resources, writing-review and editing)

ÅST (Investigation, methodology, writing-review and editing)

LJL (Investigation, methodology, writing-review and editing)

SB (Methodology, writing-review and editing)

JE (Resources, project administration, writing-review and editing)

HET (Resources, project administration, writing-review and editing)

CB (Funding acquisition, writing-review and editing)

BW (Methodology, writing-review and editing)

HC (Funding acquisition, methodology, writing-review and editing)

ØR (Funding acquisition, project administration, supervision, methodology, writing-review and editing)

SKB (Conceptualization, funding acquisition, methodology, project administration, resources, supervision, investigation, visualization, writing-original draft and editing)

## Data Availability

NA

## List of abbrevations

AN: Anorexia nervosa
BMI: Body mass index
BSFS: The Bristol Stool Scale
CIA3.0: The Clinical Impairment Assessment Questionnaire
EDE-Q6.0: The Eating Disorder Examination Questionnaire, version 6
ED: Eating disorder
EDTA: Ethylene Diamine Tetracetic Acid
EPM: Elevated plus maze
FMT: Fecal microbiota transplantation
GABA: gamma-aminobutyric acid
GAD-7: The Generalized Anxiety Disorder Assessment
GBA: The gut-brain axis
GI: Gastrointestinal
GSRS-IBS: The GI Symptom Rating Scale for IBS
HCs: Healthy controls
IBS: irritable bowel syndrome
KJ: Kilojoule
KUHR: Control and Payment of Health Reimbursements
MB: marble burying
NFC: NutriFoodCalc
NMBU: Norwegian University of Life Sciences
NORMA: Norwegian microbiota study in anorexia nervosa
Norspis: Norwegian Quality Register for the Treatment of Eating Disorders
NTNU: Norwegian University of Science and Technology
OCI-R: The Obsessive-compulsive inventory-Revised
OD: Optical density
OF: Open field
OTUs: Operational taxonomic units
PCA: Principal components analysis
PEEK: Polyether ether ketone
PHQ-9: The Patient Health Questionnaire 9
SCFAs: Short-chain fatty acids
SF-36: The 36-Item Short Form Survey
SOPs: Standard operating procedures
TP: Time point
TSD: Services for sensitive data
WP: Work pack

## Acknowledgements

We gratefully acknowledge the patients and participants for their time, commitment, and invaluable contributions to this research. We also extend our gratitude to the Norwegian user organizations for eating disorders ROS and SPISFO for their insights, contributions, and engagement in the project.

Our sincere thanks go to all the hospital units in the NORMA study, including all the medical health personnel who contribute to data collection and the administration of the study. We also appreciate the leaders of the hospital units for their support in the administration and planning of the study. Additionally, we thank the laboratory personnel who assist with the analysis and storage of biological data.

A special thanks to the healthcare personnel at the hospital units: Timea Marosi, Gro Anita Ytterstad, Bodil Hansen, Frøydis Moe, Vilde Johansen, Ane Barstad, Karina Erichsen, Hanne Tveit Horndalen, Annette Skarsbakk, May-Lis Bjørvik and Signe Haugen for their significant contributions and engagement in the project. In addition, thanks to the technicians at NMBU; Cornelia Rognstad Karlsen, Shahian Ayad Ftah and Emily Catherine Raw Kverndal

## Supporting Information

1. **S1 Supporting information**: SPIRIT checklist
2. **S2 Supporting information**: Letter of concent (in Norwegian) for AN patients
3. **S3 Supporting information**: Letter of concent (in Norwegian) for Healthy controls
4. **S4 Supporting information**: Licence Agreement GSRS use ASTRAZENECA
5. **S5 Supporting information**: Supplementary Method Description
6. **S6 Supporting information**: Ethical approvals. English translation and Norwegian original documents

